# EXPRESSION OF VARIABILITY OF THE CYP2C19*2 GENE IN PATIENTS WITH ACUTE MYOCARDIAL INFARCTION FROM A SOUTH AMERICAN HOSPITAL AND ITS FUTURE RELEVANCE IN THE INTERACTION BETWEEN GENETICS AND CARDIOLOGY

**DOI:** 10.1101/2023.08.18.23294282

**Authors:** Luis Andres Dulcey Sarmiento, Juan Sebastián Theran Leon, Jaime Gomez, Rafael Guillermo Parales Strauch, Raimondo Caltagirone, Edgar Camilo Blanco Pimiento, María Paula Ciliberti Artavia, Juan Camilo Martinez, Valentina Cabrera Peña, Maria Camila Amaya

## Abstract

**Introduction:** Some Polymorphisms of the CYP2C19 gene are associated with a decrease in the activity of the enzyme they encode, being the case of CYP2C19*2 in causing a lower generation of active metabolite of clopidogrel and therefore a low or null antiplatelet action depending on the genotype present. Antiplatelet therapy, mainly clopidogrel, is considered essential treatment in the management of acute coronary syndromes (ACS).

**Target:** The frequency of the CYPC19*2 polymorphism, identified as relevant in resistance to clopidogrel, is unknown in the population of this part of South America.

**Methods:** A descriptive, observational and cross-sectional study was designed to determine the frequency of the CYP2C19*2 allele in patients with ACS admitted to a South American hospital during the period between 2015-2016, being the first study to determine polymorphism in our population. fifty-nine adults patients diagnosed with ACS were included, 48 male (81.3%) and 11 female (18.7%), aged between 54 and 86 years. The genotype for the CYP2C19 gene was determined through the PCRRFLP (Restriction Fragments Length Polymorphism) technique from DNA extracted desde peripheral blood .

**Results:** The allelic frequency of the CYP2C19*2 polymorphism was 28.5%. Three subgroups of metabolizers were characterized : extensive (*1/*1) 40 (67.8%), intermediate (*1/*2) 17 (28.9%) and poor (*2/*2) 2 (3.3%).

**Conclusions:** This high number of carriers of the CYP2C19*2 polymorphism in the context of ACS is relevant due to its association with a lower responsiveness to clopidogrel and the possible involvement in the choice of antiplatelet therapy, for which characterization studies are required most appropriate to identify the best therapeutic strategies in our populations through pharmacogenomics.

## INTRODUCTION

Clopidogrel has been mentioned (1) and the relationship of the CYP2C19*2 allele with decreased platelet response to clopidogrel and adverse cardiovascular outcomes derived from said resistance has also been pointed out (2). The CYP2C19*2 polymorphism produces a decrease in the activity of the enzyme resulting from its encoding and therefore the antiplatelet activity of clopidogrel may be low or null depending on the genotype present in the individual.

Variations in the antiplatelet activity of clopidogrel are partly related to the ability to generate its active substance and the ability to bind to platelet receptors. In relation to this last point, variations in the P2Y12 receptor and in the generation of the active metabolite have been described; It depends firstly on its intestinal absorption, limited by some ABCB1 gene polymorphisms, and secondly, on the variability of its metabolism by the P450 enzymatic system, where CYP gene polymorphisms are involved, which produces changes in the expression or activity of the isoenzymes.

The last case is directly related to the generation of the active principle, since clopidogrel is administered as a pro-drug and its activation requires two successive oxidation reactions carried out by various enzymes of the cytochrome p450 system, which metabolize in the Liver most of the drugs used in clinical practice.

Although several enzymes of this system participate in the conversion to the active metabolite, the CYP2C19 enzyme has been most studied because, in addition to metabolizing around 45% of clopidogrel in the two oxidation reactions, it has been shown that genetic polymorphisms of the CYP2C19 gene are associated with changes in the activity of this enzyme and in the antiplatelet response to clopidogrel in patients receiving this medication (4-7). Mononucleotide polymorphisms, called SNP type (Single Nucleotide Polymorphism) of CYP2C19 produce an enzyme with no activity; these are CYP2C19*2 and CYP2C19*3. The CYP2C19*2 polymorphism consists of a guanine to adenine change at position 681 of exon 5 that leads to an alternative splicing defect. CYP2C19*3 is due to the G>A change at position 636 of exon 4, which generates a stop codon, and with it a truncated protein.

Depending on the allele present in the polymorphisms that have been described in previous studies, the enzymatic activity of this cytochrome can be normal, reduced or increased (8-11). According to the genotype, individuals can be classified into three groups, extensive metabolizers (*1/*1), intermediate (*1/*2 or *1/*3) and poor (*2/*2, *2 /*3, *3/*3) (13). The *2 allele has been mostly associated with decreased enzymatic activity and response to clopidogrel, observing a gene dose effect, where homozygous *2/*2 patients have significantly higher platelet aggregation than heterozygous patients *1 /*2 (5,12).

Among the studies published for South American mestizo populations, the one carried out in Bolivia is mentioned, in a group of 778 healthy volunteers, where they report a frequency of genotypes *1/*2 of 7.8 % and for *2/*2 of 1 % (14). The frequency of this allele for different Colombian populations has also been described, finding 15.3% for the *1/*2 genotype and 1.1% for *2/*2, in 189 subjects, also evaluating the implication of this polymorphism with treatment with proton pump inhibitors (15). Likewise, the frequency of the *1/*2 and *2/*2 genotypes has already been determined in 346 North American subjects of Mexican descent, where an incidence of 9.7 % and 3.2 % was found, respectively (16). In 2011, a frequency between 11% and 17.4% was described for the *1/*2 genotype, in different ethnic groups of different populations in Brazil (17).

In a meta-analysis published in 2010, 9 studies were grouped together, totaling 9,685 patients (54.5%) with a diagnosis of acute coronary syndrome (ACS), where the primary end point of cardiovascular death, myocardial infarction, and cerebrovascular event occurred in 863 patients. Of the patients who presented the primary end event, 71.5% were extensive metabolizers, 26.3% were intermediate metabolizers, and 2.2% poor metabolizers. Observing a significant increased risk in the primary endpoint compound in poor metabolizers, when compared to extensive metabolizers (p=0.01) (18).

The incidence of the CYPC19*2 polymorphism, which is related to clopidogrel resistance and adverse cardiovascular events, is unknown in the Venezuelan population. This is the first study carried out to determine the frequency of the CYP2C19*2 allele in patients with ACS in our country, for which patients admitted to the Hospital Universitario de los Andes in Merida Venezuela during the period between 2015-2016 with the diagnosis were included. of SCA.

## METHODS

### POPULATION AND SAMPLE

A descriptive, observational, cross-sectional study of association was elaborated, obtaining samples from patients with ancestry of Venezuelan origin in 3 generations prior to that of the participants in the study. These patients were admitted to Hospital Universitario de los Andes in Merida, Venezuela with a diagnosis of ACS. Each individual was given information about the research project and after signing the informed consent, a peripheral venous blood sample (between 5 and 15 mL) was obtained. The diagnosis of ACS was based on clinical, electrocardiographic or markers of myocardial necrosis (presence of at least 2 criteria) or angiographic findings. The included patients were classified into the different types of ACS (with ST elevation and without ST elevation). Those patients who did not meet the 3 previous Venezuelan generations, the criteria for the diagnosis of ACS and those who had a history of hepatic, autoimmune or neoplastic diseases were excluded. The collection of the samples was carried out from the beginning of 2015 to the end of 2016.

### DETERMINATION OF THE POLYMORPHISM (681G>A OF THE CYP2C19 GENE)

The samples were processed in the human genetics laboratory of the University Hospital of the University of the Andes Merida in Venezuela, using the PCR technique (Polymerase Chain Reaction)-RFLP (Restriction Fragments Length Polymorphism). Amplification of the 169 base pair product was performed using the oligomers 5’AATTACAACCAGAGCTTGGC-3’ and 5’-TATCACTTTCCATAAAAGCAAG-3’. The PCR products were digested with the restriction enzyme SmaI, whose recognition sequence is CCCGGG. Allele assignment was made by direct observation on 6% polyacrylamide gels for the presence or absence of restriction fragments. For the G allele (*1) two fragments are generated, one of 120 and another of 49 base pairs. In the presence of the A allele (*2), the substitution of the guanine in position 681 by an adenine eliminates the recognition site of the enzyme, and the amplified product is not cut.

### STATISTIC ANALYSIS

The construction of the database and the edition of the tables were carried out with the Microsoft Office 2010 Excel Program and the statistical analysis was carried out with the SPSS Statistical Package (Statistical package for the Social Sciences) version 22. The statistical analysis included the calculation of the frequencies for the group of carriers and non-carriers, separately. Contingency tables were constructed and the chi-square (χ2) statistic was calculated to assess the presence/absence of the *2 allele of the CYP2C19*2 polymorphism (carriers and non-carriers). For all the statistical tests carried out, values of p < 0.05 were considered significant.

## RESULTS

Fifty-nine adult patients diagnosed with ACS were included, 48 male (81.3%) and 11 female (18.7%), aged between 54 and 86 years. The distribution according to the diagnoses of ACS with ST elevation: 27 patients (45.77%) and ACS without ST elevation: 32 patients (54.23%) data presented in (Table 1).

**Table 1.** Distribution by gender, type of coronary event and age of the sample.

|  |  |
| --- | --- |
| Total sample | 59 patients |
| Gender | 48 male (81.3%) and 11 female (18.7%) |
| Ages included | 54 and 86 years |
| Heart attack type | 27 patients (45.77%) and ACS without ST elevation: 32 patients (54.23%) |

Three subgroups of patients were categorized according to the genotype present: extensive metabolizers (genotype *1/*1) 42 patients (71.18%), intermediate metabolizers (genotype *1/*2) 12 patients (20.33%), and metabolizers poor (*2/*2) 5 patients (8.49%); represented in (Table 2).

**Table 2.** Characteristics of the metabolizing genotype of the patients in the sample.

| Metabolizing profile | Values |
| --- | --- |
| Extensive metabolizers (*1/*1 genotype) | 42 patients (71.18%) |
| Intermediate metabolizers (*1/*2 genotype) | 12 patients (20.33%) |
| Poor metabolizers (*2/*2) | 5 patients (8.49%) |

In addition, the genotypic frequency for the CYPC19*2 polymorphism was evaluated according to the diagnosis, finding that in patients with ACS with ST elevation it was 40.7% (n=11/27) and in patients with ACS without ST elevation. ST 37.5% (n=12/32) (Table 3).

**Table 3.** Genotype frequency for the CYPC19*2 polymorphism according to the diagnosis of myocardial infarction with and without ST elevation.

| Genotypic frequency for CYP2C19*2 polymorphism | Values |
| --- | --- |
| With ST elevation | 40.7% (n=11/27) |
| ACS without elevation | TS 37.5% (n=12/32) |

## DISCUSSION

ACS are considered the leading cause of mortality worldwide, leading with 12.2% in the report of the World Health Organization in 2004 (18). Antiplatelet therapy (aspirin and clopidogrel) is part of the treatment indicated for the management of this pathology; likewise, it has been shown that the pharmacodynamic response to clopidogrel has substantial variability among these patients (7). This was confirmed when the condition called resistance to clopidogrel was identified and the first study that associated the CYP2C19*2 allele with decreased platelet response to clopidogrel was reported, evidencing adverse cardiovascular outcomes (1-3).

In our study we found that 42 patients (71.18%) were extensive metabolizers, intermediate metabolizers (*1/*2 genotype) corresponded to 12 patients (20.33%) and poor metabolizers (*2/*2) to 5. patients (8.49%) According to existing studies, the last 2 groups of patients would have increased platelet activity despite receiving clopidogrel therapy, probably due to a low response to the drug (7). The frequency of carriers of the *2 allele found in patients with ACS with ST elevation was 40.7% (n=11/27) and in patients with ACS without ST elevation 37.5% (n=12/27). 32), this was compared with that reported in other South American mestizo populations, composed mainly of healthy individuals; Observing a higher trend in the frequency of the allele in our population and with respect to the frequency of poor metabolizers in our data, it is similar to the other South American populations, except for the North American population of Mexican descent and of different ethnic groups from Brazil, which is higher. (14-17).

Although some studies suggest the relevance of carrying out tests to determine platelet aggregability in patients diagnosed with ACS and receiving treatment with clopidogrel in order to make treatment adjustments or change to another antiplatelet drug (19), currently there is no clear indication in relation to this indication (20). On the other hand, there are currently other therapeutic alternatives such as Prasugrel, which is a new derivative of thienopyridines and Ticagrelor, a P2Y12 receptor antagonist, which are more potent and effective than clopidogrel, suggesting no influence of variability due to the CYP2C19*2 polymorphism since this isoenzyme plays a minor role in its metabolism.

## CONCLUSIONS

The second study designed and conducted in the Venezuelan population where the frequency of the CYP2C19*2 polymorphism in patients with ACS is described is described. The high number of carriers of the CYP2C19*2 polymorphism, in the context of ACS and the possible altered response to clopidogrel, is relevant due to the possible implication it has in the correct choice of antiplatelet therapy. It is necessary to carry out studies that make it possible to relate the presence of the polymorphism and its association with adverse cardiovascular events, in order to identify if this appreciation is a determining factor in the prognosis of patients at the level of our latitudes.

## Data Availability

All data produced in the present work are contained in the manuscript

## Notes

### Competing Interest Statement

The authors have declared no competing interest.

### Funding Statement

This study did not receive any funding

### Author Declarations

Los Andes University Internal Medicine Ethics Committee.

